# A computational decision-support approach for personalised care in youth mental health: A pilot feasibility study protocol

**DOI:** 10.64898/2026.05.12.26353058

**Authors:** Frank Iorfino, Ashlee Turner, Mathew Varidel, Zsofi de Haan, Anna Roberts, Tianyi Zhang, Victor An, Sam Huntley, Roman Marchant, Jacob J Crouse, Sally Cripps, Sarah Barakat, Sarah Maguire, Dominic Oliver, Elizabeth M Scott, Louise Thornton, Jo Robinson, Haley M LaMonica, Ian B Hickie

**Affiliations:** Brain and Mind Centre, The University of Sydney, Sydney NSW Australia; Human Technology Institute, University of Technology, Sydney NSW Australia; School of Mathematical and Physical Sciences, University of Technology, Sydney NSW Australia; InsideOut Institute for Eating Disorders, The University of Sydney and Sydney Local Health District, Sydney NSW Australia; Department of Psychiatry, University of Oxford, Oxford UK; NIHR Oxford Health Biomedical Research Centre, Oxford UK; School of Medicine and Public Health, The University of Newcastle, Newcastle NSW Australia; The Matilda Centre for Research in Mental Health and Substance Use, The University of Sydney, Sydney NSW Australia; Orygen, The National Centre of Excellence in Youth Mental Health, Parkville VIC Australia; Centre for Youth Mental Health, The University of Melbourne, Parkville VIC Australia

**Author notes:** **Correspondence:** A/Prof Frank Iorfino, Level 5, 1 King St, Newtown NSW 2042. joint first author (Frank Iorfino, Ashlee Turner). joint senior author (Haley M LaMonica, Ian B Hickie). **Competing interests:** IBH is the Co-Director, Health and Policy at the Brain and Mind Centre (BMC), University of Sydney, which operates an early-intervention youth service at Camperdown under contract to headspace; has previously led community-based and projects supported by the pharmaceutical industry focussed on the identification and better management of anxiety and depression (Wyeth, Elil Lily, Sevier, Pfizer, AstraZeneca, Janssen, Cilag); and is the Chief Scientific Advisor to, and a 3.2% equity shareholder in, Innowell Pty Ltd, which aims to transform mental health services through the use of innovative technologies. EMS is Principal Research Fellow at the Brain and Mind Centre, The University of Sydney; Discipline Leader of Adult Mental Health, School of Medicine, University of Notre Dame; a Consultant Psychiatrist and was the Medical Director, Young Adult Mental Health Unit, St Vincent’s Hospital (Darlinghurst) until January 2021. She has received honoraria for educational seminars related to the clinical management of depressive disorders supported by Servier, Janssen and Eli-Lilly pharmaceuticals; participated in a national advisory board for the antidepressant compound Pristiq (manufactured by Pfizer); and was the National Coordinator of an antidepressant trial sponsored by Servier. DO has received consultancy fees from Google DeepMind outside of the current study.

**Keywords:** clinical decision support, mental health care, causal inference, co-design, feasibility study

## Abstract

**Introduction:** Youth mental health presentations are largely heterogenous, making it difficult to match individuals to the most appropriate interventions. Personalised, measurement-based care has the potential to improve clinical decision-making and support shared decision-making, but remains challenging to implement in routine practice. Advances in digital monitoring and causal modelling offer new opportunities to identify individual-level processes driving mental health difficulties and to generate personalised decision-support. This pilot study aims to evaluate the feasibility and acceptability of the Minding Your Mind computational decision-support approach, a newly developed approach integrating routine outcome monitoring, individual-level causal modelling, and personalised feedback to support shared decision-making between young people and their clinicians.

**Methods and analysis:** The study involves two phases. Phase 1 will recruit young people aged 15-25 years and mental health clinicians to participate in workshops to co-design the decision-support approach and its implementation into routine practice. Phase 2 is a prospective, single-arm feasibility study involving young people receiving mental health care and their treating clinicians. Primary outcomes include feasibility, acceptability, appropriateness, and usability of the decision-support approach, assessed via self-report and objective process indicators. Secondary outcomes include changes in use and experiences with shared decision-making, and clinical and functional outcomes. Quantitative analyses will be primarily descriptive, with exploratory pre-post comparisons and sensitivity analyses. Qualitative interviews will explore user experiences and implementation barriers and facilitators.

**Ethics and dissemination:** This study has been approved by the Sydney Local Health District (RPAH Zone) Human Research Ethics Committee (X25-0341). All participants will provide informed consent prior to participation. Findings will be disseminated through peer-reviewed publications, conference presentations, and accessible summaries co-developed with young people with lived experience.

**Strengths and limitations:**

- The Minding Your Mind decision-support approach combines routine outcome monitoring, individual-level causal modelling, and personalised feedback to young people and their treating clinicians, enabling a novel approach to support shared decision-making in practice.
- The two-phase mixed-methods design incorporates co-design with young people and clinicians, enhancing acceptability, implementation fidelity, and relevance to real-world youth mental health care.
- Multiple sources of feasibility data are collected, including self-report measures, objective engagement metrics, and qualitative interviews, providing a comprehensive assessment of experience and outcomes.
- The single-arm pilot design and modest sample size limit causal inference and generalisability; effectiveness outcomes are exploratory and are not intended to establish efficacy.

## 1. INTRODUCTION

The peak burden of mental illness is during the major developmental periods of youth (ages ∼12-25), with 75% of adult disorders emerging before age 25 [1,2]. Mental illness is a leading cause of disease burden among youth [3] and can disrupt the transition to adulthood with profound consequences for social relationships, school and work attainment, and physical health [4,5], highlighting the need for effective intervention during this critical period.

Although the expansion of youth mental health services has generally improved access to care [6–8], finding the most effective treatment for every individual is an ongoing challenge. Mental illness in youth is markedly heterogenous, characterised by unique and complex sets of symptoms that are driven by interactions between diverse biological, social, psychological, and behavioural mechanisms [9,10]. This presents a wide-ranging set of potential treatment targets that could improve mental health outcomes. However, many current service models of youth mental health care continue to rely heavily on reactive, non-personalised, one-size-fits-all approaches based on broad diagnostic categories and population-level guidelines. Such generic approaches fail to consider the specific processes underlying illness in individual cases and many clinicians report that they are irrelevant for many young people [11]. As a result, young people frequently receive care that does not fit their needs, which reduces satisfaction and contributes to high non-response rates, disengagement from care, and illness progression [12].

Improving outcomes for young people requires a major shift towards highly personalised and measurement-based approaches to care. Such approaches have the potential to improve clinical decision-making by identifying an individual’s multidimensional needs, matching interventions more effectively to those needs, and continuously monitoring progress to guide ongoing care based on real-time feedback about what is and is not working [13]. This approach to mental health care is also inherently supportive of shared decision-making, a critical driver of high-quality care and a desired component of mental health treatment for young people [14,15], albeit one that remains seldom adopted in practice [16]. Digital technologies offer a well-situated means of supporting this shift, making it feasible to capture rich, individual-level data on symptoms, sleep, behaviours, and functioning in real time, and recent developments in recommendation systems have begun to use these data to tailor interventions to the individual [17–19]. While largely applied in the self-management space to date, these systems illustrate how individual-level data can support personalised decision-making, providing a conceptual framework for how similar models could be applied in clinical practice for more precise decision-support.

Although this is a step towards personalised care, most recommendation systems are fundamentally limited by their reliance on observed correlations between outcomes. These systems generally treat variables as independent predictors rather than modelling how they influence one another. This matters because correlation-based recommendations may direct interventions at the wrong target. For example, psychological distress and social connectedness are strongly correlated [20], and a correlational system might recommend increasing social engagement to improve mood. However, at the population level, causal analysis of mental health data has shown that psychological distress is causally upstream of social support, functioning, sleep, and physical activity; meaning that intervening on distress can produce downstream improvements across multiple domains, whereas intervening on those downstream domains would produce effects largely isolated to themselves [21]. Causal modelling is therefore necessary to identify which variables are genuine drivers of change and which are consequences, a distinction that is invisible to correlational systems but essential for personalised intervention planning.

However, applying causal modelling at the population level is insufficient for personalised recommendations, because the causal structure linking mental health domains is not uniform across individuals. Two young people with identical symptom profiles of poor sleep, low mood, and social withdrawal may have entirely different underlying causal structures requiring different interventions. Generating truly personalised recommendations therefore requires causal modelling to be applied at the individual level, learning the causal structure specific to each person from their own longitudinal data. Recent advances framed within a structural causal modelling and Bayesian decision-theoretic framework have demonstrated the feasibility of this approach at the population level, showing how causal structure can be learned from longitudinal data and used to simulate the effects of intervening of specific domains [21]. Translating this framework to individual-level modelling, and implementing it in a form that can be used by clinicians and young people in clinical practice, remains an unaddressed gap.

### 1.1 The Minding Your Mind decision-support approach

To overcome these gaps in youth mental health care, namely the need for personalised and measurement-based care and the potential for individual-level causal modelling to enable it, we have developed The Minding Your Mind decision-support approach. This approach is designed to deliver highly personalised and measurement-based care by integrating routine outcome monitoring, computational causal modelling analyses, and personalised feedback to support shared decision-making in practice (Figure 1). The model involves three components:

**Figure 1.**
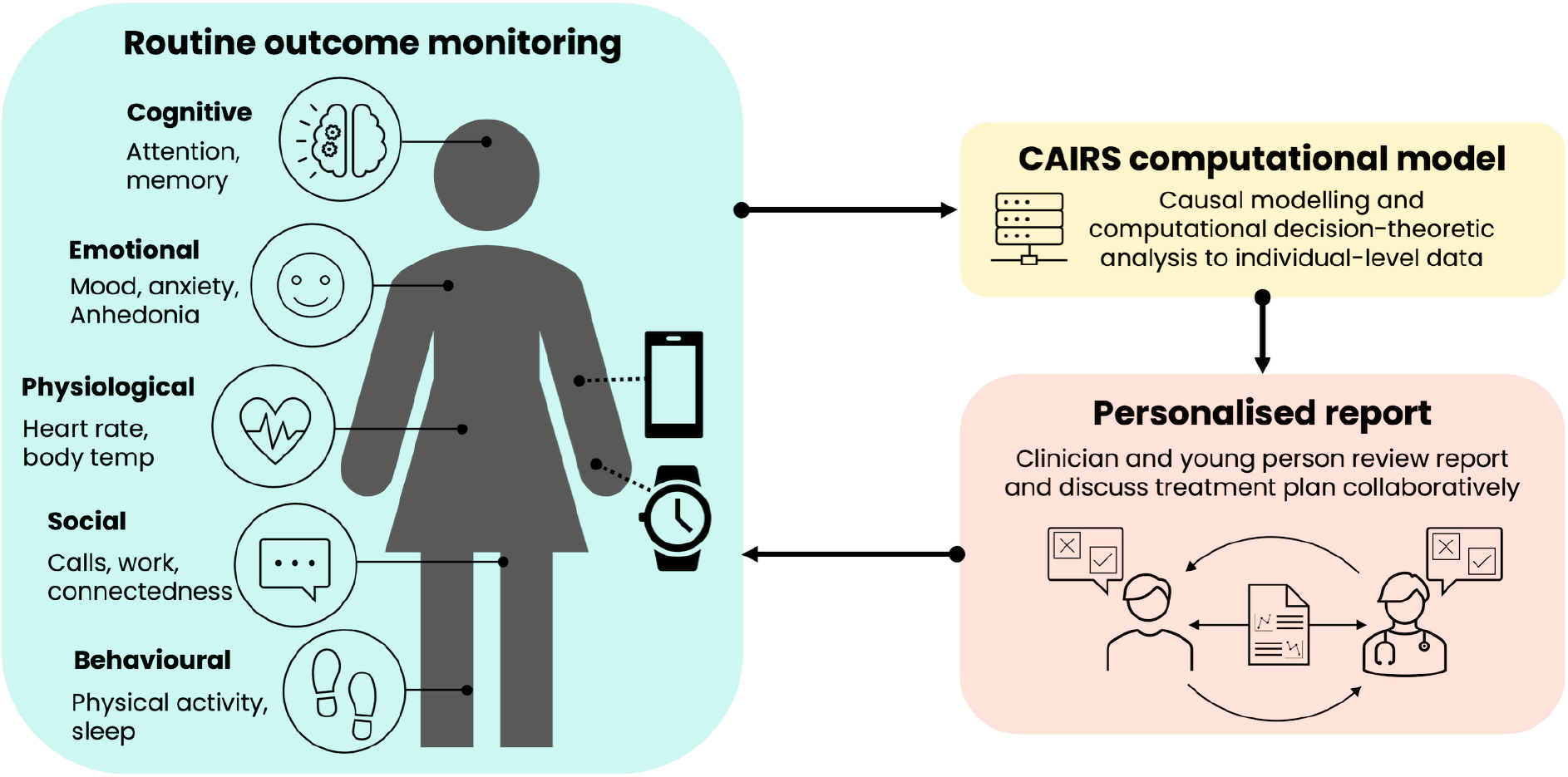
Overview of the Minding Your Mind decision-support approach

1. *Routine, high-frequency monitoring of outcomes using the Minding Your Mind smartphone app*. Building on our previous work that has established a set of differentiating factors for characterising specific processes and guiding personalised interventions [13], the *Minding Your Mind* app captures a multidimensional profile of a young person’s mental health through a combination of active (i.e., self-report surveys) and passive (i.e., device-sensed) data streams. These include sleep and circadian measures (e.g., sleep quality, duration, timing, regularity, and social jetlag), mood and emotional indicators (e.g., mood, anxiety, energy, focus, irritability), physiological and behavioural markers (e.g., resting heart rate, heart rate variability, activity levels), and social and functional participation metrics (e.g., engagement in work, study, caring, and social activities).
2. *Application of causal modelling and computational decision-theoretic analyses to longitudinal individual-level data*. Rather than examining variables in isolation or relying on correlational associations between variables, which cannot distinguish intervention targets from downstream consequences, this approach models the causal structure linking mental health domains using structural causal models. Adapting the CAIRS computational model [21] to individual-level longitudinal data, this analysis learns the causal structure to each young person—characterising how domains such as mood, sleep, behaviour, and functioning influence one another over time for that individual—and estimates uncertainty in these relationships. It also simulates idealised interventions, which models the expected effects of intervening on different domains for a given individual. By comparing expected outcomes under alternative intervention options, the model identifies the processes most likely to drive change in that individual’s mental health, while explicitly accounting for uncertainty and the interconnected nature of outcomes.
3. *Translation of causal modelling analyses into a user-centred personalised report designed to support shared decision-making and collaborative care*. Drawing on the causal modelling analyses, the report provides a shared information base, designed for both young people and clinicians simultaneously. It presents an accessible summary of the young person’s data and the inferred causal and dynamic processes underlying their current state using descriptive statistics and visualisations. It also presents ranked evidence-based intervention recommendations that reflect the processes most likely to be driving that individual’s difficulties, highlighting potential self-care goals or intervention targets aligned with the individual’s observed patterns. By presenting this information in a transparent and accessible way, the report provides a common reference point that supports young people to communicate their goals and preferences and supports clinicians to contextualise recommendations within the individual’s broader circumstances. These recommendations are therefore framed as supportive guidance rather than clinical directives, helping young people and clinicians engage in informed, collaborative discussions about priorities, preferences, and next steps in care.

The overall aim of this project is to co-design and evaluate the feasibility, acceptability, and preliminary effectiveness of the Minding Your Mind decision-support approach in youth mental health care. Specifically, the study seeks to: (1) co-design the decision-support approach, including the personalised report, to meet the needs of young people seeking mental health care and the clinicians who support them; (2) examine whether personalised reports derived from routine monitoring data and individual-level causal modelling analyses can be meaningfully integrated into clinical practice; (3) assess the acceptability and feasibility of the approach to young people and clinicians; and (4) evaluate whether the reports provide actionable, interpretable insights that enhance collaborative treatment decisions and yield preliminary indicators of clinical benefit.

#### Box 1.

Perspective by a young person with lived experience

Looking back now, I realise much of my youth was spent moving through the fog of mental illness, disconnected from my sense of self, my relationships, and the future I once imagined. What I came to mistake as normal life, and even as who I was, was in fact the persistent heaviness of undiagnosed bipolar. Although I knew something was wrong and sought help, I did not understand how deeply it was shaping every domain of my life; social, educational, psychological, and physical.

What was initially missing in my care was an integrated understanding of my mental health that went beyond symptom presentation in isolation. My care largely focused on brief snapshots in appointments, with limited insight into how sleep, stress, environment, behaviour, and biology interacted over time. Between appointments, I was left to self-manage without tools to understand patterns or track change. I did not know what “well” looked like for me, and there was little support in translating lived experience into clinically meaningful information. This gap meant I struggled to communicate what was happening internally, while clinicians often lacked the contextual data needed to fully understand my condition. As a result, I was misdiagnosed for over a decade, received treatments that were not tailored to my condition, and at times experienced interventions that worsened my trajectory. My life and self-worth fell to pieces.

Shared decision-making offers a way to bridge these gaps. To me, it means recognising that young people are not passive recipients of care, but active experts in their own experience. It means combining clinical knowledge with lived, real-time insight to make treatment collaborative rather than prescriptive. In practice, this is strengthened by routine outcome monitoring and digital health tools that capture both active and passive data, such as mood, sleep, activity, and functioning. On days when I could not engage actively, passive data reduced burden while still capturing meaningful patterns.

When care became more personalised and data-informed, it fundamentally changed my experience. It helped me see how biological, psychological, social, and environmental factors interacted to influence my symptoms, and allowed me and my clinician to adjust treatment together. Receiving personalised feedback made it easier to understand progress, communicate needs, and set goals aligned with my life circumstances. Importantly, it shifted care from something happening *to* me, to something happening *with* me, and gave me a sense of agency over something that had felt out of my control. For young people, this decision-support approach has the potential to reduce diagnostic delay, improve engagement, create a sense of empowerment and provide a clearer, more hopeful pathway through critical developmental years.

## 2. METHODS

### 2.1 Study design

This study uses a two-phase, mixed-methods design:

#### Phase 1

Participatory co-design workshops with young people and clinicians to refine the design of the Minding Your Mind personalised reports and optimise the implementation process of the decision-support model.

#### Phase 2

Prospective, single-arm, pilot feasibility study evaluating the feasibility, acceptability, and preliminary effectiveness of the Minding Your Mind decision-support model in practice.

### 2.2 PHASE 1: Implementation co-design

#### 2.2.1 Objective

The objective of Phase 1 is to co-design and refine the implementation of the Minding Your Mind decision-support model into youth mental health care. This will include co-developing the clinical protocol and processes to support the integration of the model into routine care and optimise the design of the personalised reports to support shared decision-making.

#### 2.2.2 Participants

Two participant groups will be recruited for Phase 1: young people aged 15 to 25 years (∼15) who have a history of receiving mental health care (current or previous) and clinicians currently providing mental health care to young people (∼15).

Recruitment of young people will be facilitated through multiple channels, including the research team’s existing network of youth mental health services, participating clinicians, youth mental health organisations, and advertising via the research team’s social media channels. Advertisements across these channels will include a URL to a REDCap landing page (hosted at the University of Sydney [22,23]), which will be a portal to the participant information, eligibility screening and consent process. REDCap (Research Electronic Data Capture) is a secure, web-based software platform designed to support data capture for research studies, providing: (1) an intuitive interface for validated data capture; (2) audit trails for tracking data manipulation and export procedures; (3) automated export procedures for seamless data downloads to common statistical packages; and (4) procedures for data integration and interoperability with external sources.

Clinicians will be recruited through the research team’s established clinical networks. Interested clinicians will be provided with an online link directing them to a REDCap landing page, which will be a portal to the participant information, eligibility screening and consent process.

#### 2.2.3 Consent

All individuals who are interested in participating (including young people and clinicians) will be required to provide voluntary, informed, opt-in consent. For both groups, once they enter the respective REDCap portal, they will be directed to read the participant information statement, prior to completing the eligibility screening and providing consent (if eligible).

Young people under the age of 18 years (15-to 17-year olds) will provide their own opt-in informed consent, without the need for parental involvement. This approach is supported by reseach indicating that requiring parental consent in youth health and wellbeing studies can substantially reduce participation rates and introduce systemic bias, limiting the representativeness and validity of findings [24,25]. In addition, Australian policy recognises the capacity of adolescents aged 14 years and above to make independent decisions about aspects of their health without parental oversight [26]. Information and consent materials are designed to be age-appropriate and support informed decision-making, and participation is voluntary with the option to withdraw at any time.

#### 2.2.4 Procedure

Participants will be invited to participate in co-design workshops. Workshops will be tailored specifically to young people or clinicians – i.e., young people and clinicians will not participate in the same workshops. Prior to the workshop, participants will complete a short online survey via REDCap to collect basic demographic information (e.g., age, sex). The workshops will be conducted online and facilitated by two members of the research team (including one clinician-researcher). During the workshops, participants will engage in discussions around implementation of the Minding Your Mind decision-support model and personalised reports into routine care. The purpose of the co-design workshops is to: (1) explore barriers and facilitators of shared decision-making in mental health care; (2) understand the optimal level of client data to include to meaningfully inform shared decision-making; and (3) co-create the processes supporting the implementation of the decision-support model and personalised reports into clinical practice to best support shared decision-making.

#### 2.2.5 Data collection, outcomes and analysis

The qualitative data collected through the co-design workshops will be independently coded by up to four (minimum two) investigators and analysed using established thematic analysis techniques [27]. This process will inform final refinements to the personalised report and develop the processes and protocols for implementing the decision-support model and reports into clinical practice for Phase 2.

### 2.3 PHASE 2: Pilot feasibility, acceptability and preliminary effectiveness study

#### 2.3.1 Objectives

The objective of Phase 2 is to establish the feasibility, acceptability and preliminary effectiveness of the Minding Your Mind computational decision-support model in youth mental health care.

#### 2.3.2 Participants

Two participant groups will be recruited for Phase 2: clinicians currently providing mental health care to young people and young people aged 15 to 25 years who are currently seeking or receiving care from a participating mental health clinician. Clinicians enrolled in Phase 1 form the clinician sample for Phase 2 and serve as the primary unit of recruitment of young people by raising awareness of the study to potentially eligible young people from their current caseload.

Young people are eligible if they:

1. Present with depressive symptoms, as indicated by a score of ≥6 on the self-report Quick Inventory of Depressive Symptomatology (QIDS-SR) [28]; and/or
2. Present with circadian disturbance, as indicated by a rating of ‘moderate’ or ‘high’ on the Brain and Mind Centre Sleep-Wake questionnaire; and
3. Are currently receiving individual clinical care from a participating clinician;
4. Are proficient in English; and
5. Have access to a smartphone and internet connection; and
6. Are willing and able to give online informed consent.

#### 2.3.3 Consent

Consent procedures for Phase 2 will follow those used in Phase 1. Clinicians provide a single informed consent covering participation in both Phase 1 and Phase 2 and will not be reconsented for each young person who participates. Young people recruited for Phase 2 represent a distinct cohort from those involved in Phase 1 and will provide phase-specific opt-in informed consent prior to participation.

#### 2.3.4 Procedure

An overview of the study flow is shown in Figure 2. After consenting, young people will download and complete the onboarding process for the *Minding Your Mind* app, which includes setting up a profile, setting Apple Health data sharing permissions (for iPhone users), and completing a short questionnaire. They will be asked to complete the daily check-in every day for a period of four weeks. The daily check-in has been designed to be quick and easy to complete (nine items, approximately five minutes to complete) and asks about various aspects of mood, social connectedness, and engagement with work/school (see Supplemental Material). Data from Apple Health are shared automatically with the app (if permission is granted). Participants are free to engage with the other features of the app (e.g., psychoeducation, behavioural strategies) at their own discretion. After four weeks, data from the *Minding Your Mind* app will be securely transmitted via an Application Programming Interface (API) for analysis and generation of their personalised report.

The data will be analysed using the adapted CAIRS computational model [21] that summarises relationships between mood, sleep, behaviours, and other metrics, and provides brief evidence-based recommendations for care, based on the individual’s observed data patterns (see Supplemental Material for a full list of data collected via the app and included in the modelling analysis). Once the data has been processed and the report generated, the report will be sent to both the young person and their clinician. At their next scheduled appointment, the young person and their clinician will review the report together, using it as a foundation for shared-decision making about their current needs, goals and potential care strategies (according to the processes co-designed during Phase 1). Clinical care will continue as normal and young people will be encouraged to continue to engage in monitoring through the *Minding Your Mind* app as a part of their ongoing care. For those who choose to continue monitoring their outcomes, updated reports will be sent monthly (for up to three months), and clinicians can request an updated report at other time intervals if needed.

The personalised report is designed to support clinical decision-making by providing transparent, interpretable information that can be considered alongside other clinical inputs. It does not provide diagnostic conclusions or treatment directives, and clinical judgement remains with the treating clinician. As such, it is proposed to meet criteria for exemption from regulation as a clinical decision support software as outlined in the Therapeutic Goods (Medical Devices) Regulations 2002, Australia [29]. An exemption notice process has been initiated with the Therapeutic Goods Administration and will be competed prior to commencing Phase 2. Any care plan resulting from the recommendations provided in the personalised report forms a part of standard clinical practice and is not part of the research study. All clinical decisions remain at the discretion of the treating clinician, and no data will be collected regarding what interventions were selected.

##### App engagement support

One of the key implementation risks is low adherence to the daily check-in schedule during the four-week monitoring period. Minimising gaps in daily check-in completion is critical for maximising the utility and interpretability of the personalised reports. Preliminary analyses indicate that a minimum of 14 days of consecutive daily check-ins is required to generate sufficiently reliable estimates of an individual’s mood and behaviour patterns, as well as the relationships between these variables. To support complete onboarding and sustained engagement during the active monitoring period, several complementary strategies have been implemented. First, participant’s will be provided with a welcome pack, which details each step to download the Minding Your Mind app, complete the onboarding process, and navigate the key areas they will need. Second, the Minding Your Mind app will use one-daily push notifications to prompt participants to complete their check-ins. Such in-app notifications are a simple and widely used strategy in digital health and have been shown to support engagement [30]. Third, human-support from a digital navigator will be provided as and when needed throughout the study. Digital navigators are increasingly being integrated into healthcare settings to support the use of digital health technologies and have been shown to improve digital literacy, confidence, and engagement [31,32]. At enrolment, the digital navigator will be available to provide onboarding support, including guidance on downloading the app, completing the onboarding process, and navigating key app features. They will also monitor engagement and provide ongoing support throughout the four-week monitoring period. If a participant has not completed a daily check-in for three consecutive days, the digital navigator will initiate contact to provide a gentle reminder and assist with troubleshooting any technical difficulties. Finally, participants will receive financial compensation for completing daily check-ins. Evidence from studies involving intensive data collection indicates that per-day compensation can increase willingness to participate and improve adherence to repeated data collection schedules [33,34]. Young people will receive $2 AUD for each daily check-in completed (up to $14 AUD per week) and an additional $10 AUD bonus for completing at least 85% of check-ins within a given week (i.e., ≥6 days completed). Participants can receive up to $96 AUD in total for meeting the weekly completion threshold across the four-week monitoring period. Daily check-ins completed beyond this initial four-week period will not be compensated.

#### 2.3.5 Sample size

As the primary aim of Phase 2 is to assess the feasibility and acceptability of the Minding Your Mind decision-support approach, sample size was determined in line the guidance for pilot and feasibility studies and did not involve a formal power calculation.

##### Clinician sample

The target sample size is 12 clinicians. This figure is consistent with established rules-of-thumb for pilot studies suggesting a minimum of 12 participants per group as sufficient to yield preliminary estimates of key parameters [35,36]. Accounting for an assumed dropout rate of 20% by the end of the study, 15 clinicians will be recruited. This guidance is appropriate here given the nature of the clinician-level feasibility questions: the Minding Your Mind decision-support approach places relatively low burden on clinicians. The personalised report is delivered as supplementary clinical information that can be integrated into existing consultations, rather than requiring a new procedural protocol, meaning the feasibility questions at this level are therefore less demanding to estimate than those at the young person level. Specifically, the clinician-level feasibility objectives concern whether clinicians can be recruited and retained, whether the report is perceived as useful and feasible to integrate into practice, and whether it helps them to engage in collaborative treatment planning with their clients. The continuity of clinicians across phases is intentional: having co-designed the implementation procedures in Phase 1, they bring directly relevant procedural knowledge into the pilot, which strengthens the intervention fidelity.

##### Young person sample

The target sample size is 40 young people. This sample size was selected according to published recommendations and minimum rules-of-thumb for pilot studies [37,38]. It was also guided by comparable and relevant prior pilot feasibility studies in the digital mental health field with young people [39,40]. Accounting for an assumed dropout rate of 30% by the end of the study, 58 young people will be recruited (up to 5 recruited from each participating clinician). The young person feasibility questions are more demanding than those at the clinician level and justify a larger sample size relative to the clinician group. The core feasibility questions for young people concern whether sustained daily monitoring is achievable over four weeks, whether the personalised report is comprehensible and meaningful to them, and whether it supports a sense of shared understanding with their clinician that enables them to participate more actively in decisions about their care. These outcomes span behavioural adherence, data acceptability, and patient-reported experience, and require sufficient participants to estimate each on a useful level.

#### 2.2.6 Data collection, outcomes and analysis

The primary outcomes are feasibility and acceptability, with preliminary effectiveness as an exploratory outcome. The mixed-methods study design will collect a combination of quantitative and qualitative data that will be analysed to understand the feasibility, acceptability, and preliminary effectiveness of the Minding Your Mind decision-support model in youth mental health care.

##### Quantitative

Quantitative data will be collected via online self-report surveys. Surveys will be completed by participants (young people and clinicians) at baseline (month 0) and various timepoints thereafter (month 1, 2 and 3). The schedule of assessments according to each participant group is outlined in Table 1 and a detailed description of each assessment measure is included in the Supplemental Material. To complement self-report measures, feasibility and acceptability of the decision-support model and study procedures will be assessed using a range of objective indicators (Table 2). Together, these outcomes will provide insight into the practicality, usability, and sustainability of this approach in real-world clinical settings and to inform a larger scale implementation trial in the future.

**Table 1.** Schedule of outcome domains, assessment measures and timepoints for data collection.

| Domain | Assessment | User group | Timepoint (months) |  |  |  |
| --- | --- | --- | --- | --- | --- | --- |
|  |  |  | 0 | 1 | 2 | 3 |
| *Feasibility* | FIM (feasibility) [45] | YP, MHC |  | ✓ |  |  |
| *Acceptability and appropriateness* | AIM (acceptability) [45] | YP, MHC |  | ✓ |  |  |
|  | IAM (appropriateness) [45] | YP, MHC |  | ✓ |  |  |
| *Usability* | SUS [46] | YP, MHC |  | ✓ |  |  |
| *Preliminary effectiveness* | SDM-Q-9 [47] (shared decision making) | YP | ✓ | ✓ | ✓ | ✓ |
|  | SDM-Q-Doc [48] (shared decision making) | MHC | ✓ | ✓ | ✓ | ✓ |
|  | QIDS-17-SR [28] (depressive symptoms) | YP | ✓ | ✓ | ✓ | ✓ |
|  | Sleep-wake cycle questionnaire (circadian disturbance) | YP | ✓ | ✓ | ✓ | ✓ |
|  | GAD-7 [49] (anxiety symptoms) | YP | ✓ | ✓ | ✓ | ✓ |
|  | WSAS [50] (daily functioning) | YP | ✓ | ✓ | ✓ | ✓ |
|  | AQoL-6D [51] (quality of life) | YP | ✓ | ✓ | ✓ | ✓ |

Note. YP, young person; MHC, mental health clinician; FIM, Feasibility of Intervention Measure; AIM, Acceptability of Intervention Measure; IAM, Intervention Appropriateness Measure; SUS, System Usability Scale; SDM-Q-9, Shared Decision Making Questionnaire (Patient version); SDM-Q-Doc, Shared Decision Making Questionnaire (Health Professional version); QIDS-17-SR, Quick Inventory of Depressive Symptomatology-17 item-Self-Report; GAD-7, Generalised Anxiety Disorder; WSAS, Work and Social Adjustment Scale; AQoL-6D (Assessment of Quality of Life).

**Table 2.**
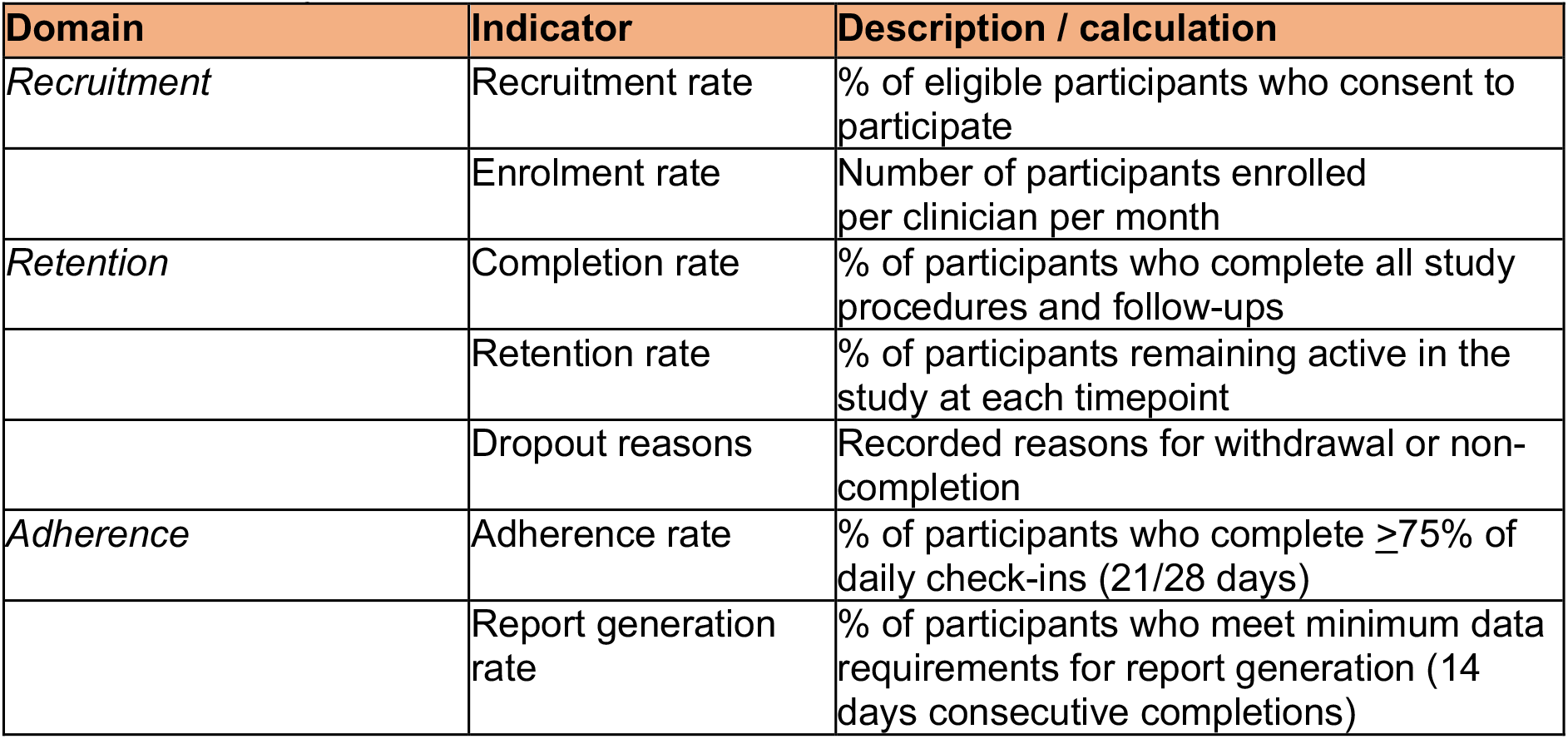
Feasibility indicators.

| Domain | Indicator | Description / calculation |
| --- | --- | --- |
| *Recruitment* | Recruitment rate | % of eligible participants who consent to participate |
|  | Enrolment rate | Number of participants enrolled per clinician per month |
| *Retention* | Completion rate | % of participants who complete all study procedures and follow-ups |
|  | Retention rate | % of participants remaining active in the study at each timepoint |
|  | Dropout reasons | Recorded reasons for withdrawal or non-completion |
| *Adherence* | Adherence rate | % of participants who complete $\geq 75\%$ of daily check-ins (21/28 days) |
|  | Report generation rate | % of participants who meet minimum data requirements for report generation (14 days consecutive completions) |

All statistical analyses will be conducted using R [41]. Descriptive data will be presented as mean ± standard deviation for continuous variables and frequencies and percentages for categorical variables. Feasibility, acceptability, appropriateness, and usability outcomes will be summarised descriptively using mean scale scores and distributions for each outcome. Within-person changes in shared decision-making and clinical and functional outcomes from baseline to follow-up will be analysed using paired samples analyses (e.g., paired samples t-tests or non-parametric equivalents). Correlational analyses will examine associations between shared decision-making indicators, and clinical and functional outcomes. Given the feasibility design, all inferential analyses are exploratory and will be interpreted accordingly.

Informative dropout is an anticipated challenge of this study, as participants with lower perceived feasibility may complete fewer daily check-ins and be less likely to complete follow-up surveys, meaning completer analysis would likely overestimate feasibility. Missing outcome data will therefore be handled used multiple imputation by chain equations (MICE) [42], with indicators of adherence (e.g., observed follow-up data, daily check-in completion rates) included as predictors in imputation models to account for the systematic relationship between engagement and perceived feasibility. Primary analyses will pool estimates across imputed datasets using a Bayesian modelling framework [43]. A delta-adjustment sensitivity analysis will be conducted to assess robustness to missing-not-at-random assumptions [44].

##### Qualitative

Qualitative data will be collected via one-on-one semi-structured interviews with participants. Clinicians and young people will be invited to participate in an interview after completing the follow-up surveys at the month one timepoint (i.e., once the young person has completed the four-week monitoring period, had their report made available to use during an appointment with their clinician, and completed the follow-up survey). Clinicians will be invited once they have had at least one of their clients meet these criteria. Interviews will be conducted online (via Zoom) by a member of the research team. The purpose of the interviews is to provide additional feedback on participant experiences using the Minding Your Mind decision-support model and personalised reports for shared decision-making. Based on Sekhon’s framework for acceptability [52], interviews will explore attitudes, burden, ethicality, perceived effectiveness, and self-efficacy in relation to the Minding Your Mind decision-support model within youth mental health care. The qualitative data collected through the interviews will be independently coded by up to four (minimum two) investigators and analysed using established thematic analysis techniques [27]. This process will develop an understanding of the key concepts relating to the implementation of the decision-support model and personalised reports into care and their ability to support shared decision-making between young people and their clinicians.

#### 2.3.7 Progression criteria

Following recommendations for feasibility study design, pre-specified progression criteria will be used to guide the decision of whether to proceed to a future trial, using a traffic light framework. Thresholds for each criterion were established a priori (Table 3). Report generation rate will be the primary feasibility threshold given its direct operational significance. Ecological momentary assessment (EMA) compliance in clinical youth populations varies with assessment frequency and duration: studies using higher-frequency protocols (e.g., six assessments per day over seven days) have reported compliance rates around 69% [53], while lower-frequency protocols (e.g., two to three assessments per day over 14-21 days) have demonstrated compliance rates of 80-90% [54,55]. Accounting for the stringency of the consecutive-days requirement, a green threshold of ≥70% is considered conservative but realistic, and is consistent with the pooled EMA compliance estimates in general child and adolescent samples (72%) [56].

**Table 3.** Proposed progression thresholds for proceeding to a future trial.

| **Outcome** | **Green limit** | **Amber limit** | **Red limit** |
| --- | --- | --- | --- |
| Report generation – *achieve at least 14 consecutive daily check-ins* | $\geq 70\%$ | 50-69% | $< 50\%$ |
| Recruitment rate – *the proportion of eligible participants who consent to participate.* | $\geq 60\%$ | 40-59% | $< 40\%$ |
| Retention rate – *the proportion of enrolled participants remaining active at the one-month timepoint.* | $\geq 75\%$ | 55-74% | $< 55\%$ |
| Completion rate – *the proportion of enrolled participants who complete all study procedures and follow-up assessments.* | $\geq 65\%$ | 45-64% | $< 45\%$ |
| Acceptability – *mean acceptability score on the AIM (total sample)* | $\geq 4$ | 3-3.9 | $< 2$ |

The pilot study will proceed to a full trial if ≥3 of these criteria are met in the green zone. In the event that criteria are partially met at this threshold, modifications to the design and/or decision-support approach will be undertaken before proceeding. Failure to meet this threshold, or any criterion falling in the red zone, indicates the study should not proceed without substantial redesign [57].

### 2.4 Data management

Electronic consent will be obtained via the University of Sydney’s instance of the secure online data capture platform REDCap. Survey data will also be collected via REDCap using a secondary project system to ensure identifying information and research data are stored separately.

Audio recordings from co-design workshops and semi-structured interviews will be immediately transferred to a secure, password-protect, enterprise-grade server at the University of Sydney (the Research Data Store [RDS]). Audio recordings will be transcribed using an external third-party service (Otter.ai) and all files (audio recordings and text-based transcriptions) will be deleted from the Otter.ai platform immediately after transcription. Once the transcription process is complete, audio recordings will be permanently deleted from the RDS. Text-based transcriptions will be transferred and stored on the RDS.

On completion of the project, all research data will be stored on the University of Sydney’s RDS. All study data will be de-identified, coded numerically, password-protected, and accessible only to named researchers on the project.

All data entered into the *Minding Your Mind* app is subject to the *Minding Your Mind* privacy policy. Contact information (i.e., email address needed to create an account for the app) and user generated data (i.e., survey responses and passive data) is encrypted and stored securely in separate password protected files on Mongo DB in its Sydney data centres. The data centres are SOC II/III certified and are highly secured, maintained and monitored. The app infrastructure, policies, and processes are FERPA and HIPAA compliant. After the four-week monitoring period, data will be securely transmitted via an API to the University of Sydney’s instance of Amazon Web Services (AWS) for processing. Data are temporarily held in AWS for analysis and report generation and deleted immediately after the report is generated. Once the report has been sent to the clinician and young person, the reports are deleted and not stored.

### 2.5 Study duration

The study is expected to run for two years, starting from April 2026.

## 3. PATIENT AND PUBLIC INVOLVEMENT

Young people with lived experience and mental health clinicians have been involved across all stages of developing this research, with the aim of ensuring the study and its procedures are acceptable, meaningful, and feasible for the people they are designed to serve.

To date, involvement has included contribution to the project proposal for the funding application, with a young person with lived experience (ZdH) a named investigator on the successful application. Several named investigators (SB, SM, EMS, HML, IBH) are mental health clinicians whose experience working with young people and within health services helped shape the study design and direction. The final study protocol was reviewed and approved by young people with lived experience, including salaried lived experience researchers on the Brain and Mind Centre (BMC) research team and the BMC’s Lived Experience Working Group (LEWG) [58].

Initial versions of the personalised report were designed and iterated collaboratively with young people with lived experience. Version 1 of the report was presented to the LEWG in a facilitated workshop, for which members were remunerated for their time. Feedback was used to iterate to version 2, which was subsequently re-reviewed to confirm whether initial concerns were addressed and identify any further issues or improvements needed. Lived experience researchers on the BMC research team also provided direct feedback on the developing versions of the report. A detailed log of all feedback was maintained alongside documentation of how each item was incorporated, or where it could not be, the rationale for this decision, which was fed back to contributors for transparency. Feedback received from the LEWG and lived experience researchers resulted in meaningful change to the report design, including the use of visual presentation of data over numerical summaries, appropriate use of colour, and restructuring the report to start with higher-level insights and recommendations before presenting more detailed data. As part of Phase 1, young people and clinicians will contribute to final refinements to the report and co-design its integration into clinical workflows prior to implementation. The complete process for designing the reports from the initial theoretical approach to the final design will be highlighted in a subsequent publication.

Young people with lived experience (including salaried researchers and the LEWG) will be involved in various capacities across the duration of the project, including advertising of the study through youth-focussed channels, supporting the interpretation of findings, and knowledge dissemination.

## Data Availability

No datasets were generated or analysed during this study, as this is a study protocol.

## Acknowledgements

The authors would like to acknowledge, in advance, the young people and clinicians who will participate in this study.

## 4. ETHICS AND DISSEMINATION

The study protocol has been approved by the Sydney Local Health District (RPAH Zone) Human Research Ethics Committee (X25-0341).

Findings from this study will be disseminated to academic, clinical, and community audiences, with a particular emphasis on ensuring outputs are accessible and meaningful to young people. Results will be submitted for publication in peer-reviewed journals, with open access publication prioritised to maximise reach. Findings will be presented at national and international conferences relevant to youth mental health, digital health, and implementation science. Plain language summaries tailored to clinicians will be developed to support translation of findings into practice, and results will be disseminated to partner youth mental health services as a topic of interest related to service development. A policy brief will be prepared summarising key implementations for youth mental health service design and the implementation of personalised, data-driven care models. To ensure the findings are disseminated to young people in an accessible, relevant, and appropriately framed manner, lived experience researchers on the BMC research team and the LEWG will be involved in interpreting findings and shaping how results are communicated. Plain language summaries written specifically for young people will be developed collaboratively with lived experience contributors and made freely available through the Minding Your Mind website. Short-form video content summarising key findings will be produced in collaboration with young people and disseminated through the Lived Experience Lab Instagram account and shared across partner youth networks. This channel has been selected for its established reach among young people and its grounding in lived experience perspectives. Additional dissemination through broader youth-focussed networks and partner organisations will be pursued to maximise community reach beyond the immediate research team.

