## Supplementary material for "A computational decision-support approach for personalised care in youth mental health: A pilot feasibility study protocol"

1. ***Minding Your Mind app – daily check-in items***
2. ***Minding Your Mind app – Apple Health data points***
3. ***Detailed description of Phase 2 self-report surveys***

***Minding Your Mind daily check-in items***

1. How happy vs. sad do you feel right now?

| 1 *Very cheerful / happy* | 2 | 3 | 4 | 5 | 6 | 7 *Very sad / depressed / unhappy* |
|---|---|---|---|---|---|---|

2. How calm or relaxed vs. nervous/anxious do you feel right now?

| 1 *Very relaxed / calm* | 2 | 3 | 4 | 5 | 6 | 7 *Very nervous / anxious* |
|---|---|---|---|---|---|---|

3. How tired vs. energetic do you feel right now?

| 1 *Very tired / sluggish* | 2 | 3 | 4 | 5 | 6 | 7 *Very energetic / lively* |
|---|---|---|---|---|---|---|

4. How well can you concentrate or focus right now?

| 1 *Very focused / attentive* | 2 | 3 | 4 | 5 | 6 | 7 *Very unfocused / distracted* |
|---|---|---|---|---|---|---|

5. How irritable or easily angered do you feel right now?

| 1 *Not at all irritable / angry* | 2 | 3 | 4 | 5 | 6 | 7 *Very irritable / angry* |
|---|---|---|---|---|---|---|

6. [Last night] I slept well

| 1 *Not at all* | 2 | 3 | 4 | 5 | 6 | 7 *Very much* |
|---|---|---|---|---|---|---|

7. How close to vs disconnected from others did you feel today?1 (Not at all irritable/angry)

| 1 *Not connected at all* | 2 | 3 | 4 | 5 | 6 | 7 *Very connected* |
|---|---|---|---|---|---|---|

8. How much time did you spend in meaningful social interactions today?

- 0 minutes
- 1-30 minutes
- 31-59 minutes
- 1-2 hours
- 2-4 hours
- >5 hours

9. How much time did you spend engaged in education, study, work, caring for others or volunteering?

- Today was my day off from these activities
- 0
- 1 hour
- 2 hours
- 3 hours

- 4 hours
- 5 hours
- 6 hours
- >7 hours

***Minding Your Mind Apple Health data points***

| Category | Data element |
|---|---|
| Activity | Active energy |
| Activity | Resting energy |
| Activity | Exercise minutes |
| Activity | Stand hours |
| Activity | Steps |
| Activity | Walking + running distance |
| Activity | Cycling distance |
| Activity | Swimming distance |
| Activity | Flights climbed |
| Activity | Workouts |
| Body measurement | Weight |
| Body measurement | Height |
| Heart | Heart rate |
| Heart | Heart rate variability |
| Heart | Resting heart rate |
| Heart | Walking heart rate |
| Mindfulness | Mindful minutes |
| Sleep | Sleep duration |
| Sleep | Time in bed |
| Sleep | Sleep time |
| Sleep | Wake time |
| Sleep | Wrist temperature |
| Vitals | Respiratory rate |

***Detailed description of Phase 2 self-report surveys***

***Feasibility of Intervention Measure (FIM):*** The FIM is a brief, self-report tool designed to assess the perceived practicality and implementability of a healthcare intervention from the perspective of stakeholders, including clinicians and patients. The FIM consists of four items, each rated on a 5-point Likert scale ranging from 1 ("completely disagree") to 5 ("completely agree"), with higher scores indicating greater perceived feasibility.

***Acceptability of Intervention Measure (AIM):*** The AIM is a brief, self-report tool designed to assess the perceived acceptability of a healthcare intervention from the perspective of stakeholders, including clinicians and patients. The AIM consists of four items, each rated on a 5-point Likert scale ranging from 1 ("completely disagree") to 5 ("completely agree"), with higher scores indicating greater perceived acceptability.

***Intervention Appropriateness Measure (IAM):*** The IAM is a brief, self-report tool designed to assess the perceived fit, relevance, or suitability of a healthcare intervention for a particular setting or population, from the perspective of stakeholders including clinicians and patients. The IAM consists of four items, each rated on a 5-point Likert scale ranging from 1 ("completely disagree" to 5 ("completely agree"), with higher scores indicating greater perceived appropriateness.

***System Usability Scale (SUS):*** The SUS is a widely used, validated self-report questionnaire designed to assess the usability and user experience of digital product/system/intervention. The SUS consists of 10 items; each rated on a 5-point Likert scale ranging from 1 ("strongly disagree") to 5 ("strongly agree"). Individual item scores are summed and multiplied by 2.5 to yield a total scale ranging from 0 to 100, with higher scores indicating better usability.

***Quick Inventory of Depressive Symptomatology, 16-item, Self-Report (QIDS-16-SR):*** The QIDS is a validated self-report tool designed to assess the severity of depressive symptoms across nine symptom domains (sleep, sad mood, appetite/weight, concentration/decision making, self-view, thoughts of death or suicide, general interest, energy levels, and restlessness/agitation). Each symptom domain is scored from 0 to 3 and the total score ranges from 0 to 27, with higher scores indicating greater depressive symptom severity.

***Sleep-wake cycle questionnaire:*** This questionnaire includes six questions that ask about bedtimes during weekdays and weekends, waking time during weekdays and weekends, hours of sleep, and feelings when waking up.

***Generalised Anxiety Disorder-7 (GAD7):*** The GAD-7 is a validated self-report tool designed to assess the severity of generalised anxiety symptoms. The scale consists of seven items, each reflecting a key symptom of anxiety (e.g., nervousness, uncontrollable worry, restlessness). Each item is rated based on how often each symptom has been experienced in the past two weeks using a 4-point Likert scale: 0 ("not at all") to 3 ("nearly every day"). The total score ranges from 0 to 21, with higher scores indicating greater anxiety severity. Standard cut-offs are commonly used to categorise anxiety levels as minimal (total score 0-4), mild (total score 5-9), moderate (total score 10-14), and severe (total score 15-21).

***Work and Social Adjustment Scale (WSAS):*** The WSAS is a brief, self-report tool designed to assess the functional impairment assessment with a mental health condition or other health problem. It includes five items, each assessing the impact of a problem on a different domain of daily functioning: work, home management, social leisure, private leisure, and close relationships. Each item is rated on an 8-point Likert scale from 0 ("not at all impaired") to 8 ("very severely impaired"), with a total score ranging from 0 to 40 and higher scores indicating greater functional impairment.

***Assessment of Quality of Life, 6 Domains (AQoL-6D):*** The AQoL-6D is a validated self-report tool designed to measure health-related quality of life across multiple domains. It includes 20 items covering six dimensions: independent living, relationships, mental health, coping, pain, and senses. Each item is scored using multi-level response options, and responses are combined according to the scoring algorithm to produce dimension scores and a single utility index ranging from 0 (worst possible quality of life) to 1 (full quality of life).

***SDM-Q-9:*** The SDM-Q-9 is a validated self-report instrument designed to assess the extent to which patients perceive that shared decision-making has occurred during a clinical consultation. The questionnaire includes nine items, each reflecting a specific step in the shared decision-making process. Each item is rated on a 6-point Likert scale ranging from 0 ("completely disagree") to 5 ("completely agree"), yielding a raw total score between 0 and 45, with higher scores indicating a greater level of shared decision-making from the patient's perspective.

***SDM-Q-Doc:*** The SDM-Q-Doc is the clinician-reported counterpart to the SDM-Q-9, designed to assess healthcare professionals' perceptions of the shared decision-making process. The questionnaire includes nine items that parallel those of the SDM-Q-9, each representing a key element of shared decision-making. Each item is rated on a 6-point Likert scale ranging from 0 ("completely disagree") to 5 ("completely agree"), yielding a raw total score between 0 and 45, with higher scores indicating a greater level of shared decision-making from the patient's perspective.
